# COVID-19 cluster size and transmission rates in schools from crowdsourced case reports

**DOI:** 10.1101/2021.12.07.21267381

**Authors:** Paul Tupper, Shraddha Pai, COVID Schools Canada, Caroline Colijn

## Abstract

The role of schools in the spread of the COVID-19 pandemic is controversial, with some claiming they are an important driver of the pandemic and others arguing that transmission in schools is negligible. School cluster reports that have been collected in various jurisdictions are a source of data about transmission in schools. These reports consist of the name of a school, a date, and the number of students known to be infected. We provide a simple model for the frequency and size of clusters in this data, based on random arrivals of index cases at schools who then infect their classmates with a highly variable rate, fitting the overdispersion evident in the data. We fit our model to reports from four Canadian provinces, providing estimates of mean and dispersion for cluster size, whilst factoring in imperfect ascertainment. Our parameter estimates are robust to variations in ascertainment fraction. We use these estimates in two ways: i) to explore how uneven the distribution of cases is among different clusters in different jurisdictions (that is, what fraction of cases are in the 20% largest clusters), and ii) to determine the distribution of instantaneous transmission rate *β* among different index cases. We show how these latter distributions can be used in simulations of school transmission where we explore the effect of different interventions, in the context of highly variable transmission rates.

## Introduction

In the management of the COVID-19 pandemic an important consideration is the role of children and in particular schools. In most jurisdictions rates of SARS-CoV-2 infection among children are similar to those in the adult population (*10*). But severity is much lower in children; the infection fatality rate (IFR) of COVID for at age 10 was estimated to be 0.002% versus an IFR of 0.01% at age 25, and 0.4% at age 55, for the original SARS-CoV-2 virus present in 2020 (*26*). Cases are more often asymptomatic among children, less likely to require hospitalization and ICU care (*10*), and less likely to be classified as long COVID (*37*). On the other hand, MIS-C is a serious condition sometimes resulting from SARS-CoV-2 infection (*7*), and myocarditis happens more frequently as a side effect of infection among younger individuals (*34*).

Jurisdictions have had to make a choice between closing schools, with all the attendant social, economic, and psychological costs (*11*), and leaving schools open, allowing possible transmission of SARS-CoV-2 in that setting (*10*). The direct downside of transmission in schools if it occurs is that children may be infected there, risking the low but non-negligible harms of COVID-19 in that age range, but also adult teachers and staff are put at risk. Transmission in schools may also contribute to overall community transmission, indirectly jeopardizing more vulnerable individuals (*44*). As a concrete example, if a child contracts SARS-CoV-2 at school, they may then go on to transmit it to an elderly relative they live with, for whom the consequences are more severe (*23*). Estimating the magnitude of these two kinds of harm and making the decision as to what choice to make involves many sources of uncertainty and value judgements, which helps explain why different jurisdictions have taken different approaches (*20*). In some jurisdictions schools were open for the 2020 to 2021 school year, though many measures were put into place in order to reduce the risk of SARS-CoV-2 transmission (*30*). Measures include cohorting, staggered entrance and exit times, masks, improvements in ventilation, extra sanitization measures. In other jurisdictions schools were closed for large portions of the year (*32*).

Studies that have looked at the effect of school closures on the overall rate of SARS-CoV-2 transmission find mixed results: some find substantial reduction in community transmission when schools are closed, and others small or no effect (*13,44*). Given that schools involve many children all sharing a room for many hours a day, it may be surprising that there is not a clearer evidence of significant transmission in schools. One explanation is that children may be less likely to transmit SARS-CoV-2 to each other, either by being less infectious or by being less susceptible (*16, 41*). But transmission in schools does occur, and it’s worthwhile to estimate the magnitude and characterize the variation in it.

One source of evidence for transmission in schools are school exposure reports. Throughout the pandemic organizations have collected data submitted by volunteers about COVID cases in schools, and such data has subsequently been published online (*4, 6*). Data consists of reports of exposures or clusters in schools, either submitted by parents, or determined from reading newspaper reports. Several such websites exist, though many ceased due to excessive workload after the 2020–21 school year. In some jurisdictions there are also similar sources of data provided by local government (*18,31*) or Public Health Agencies (*21, 22*).

Here we propose a simple model of transmission in schools, and we use these data on cluster sizes to estimate parameters of the model for four Canadian provinces. Our model allows for heterogeneity in transmission rate, which is able to capture the considerable variability in the sizes of the clusters, with most exposures leading to no further cases (and so a cluster of size 1) but with few having a large number of cases (*39*). We estimate the mean and overdispersion parameters for different jurisdictions. We then use our parameter estimates in a couple of ways: firstly, we explore the overdispersion of cluster sizes in different jurisdictions, giving estimates of what fraction of all cases are in the 20% largest of all clusters. Secondly, we can obtain an estimate of the distribution of the transmission rate *β*, the rate at which a single infected individual infects a susceptible person when they are in contact. This parameter, in turn, could be used to simulate school transmission and explore the impacts of interventions (*40*) as we explore for some parameter choices. In the Supplementary Information we perform a similar analysis for eight US states, where only substantially less complete data sets were available.

Finally, two important changes have occurred in 2021 that we expect to impact cluster sizes in schools. On the one hand, in many jurisdictions, large portions of children aged 5 and up have been vaccinated with the Pfizer/BioNTech vaccine (*38*). According to the extent to which the vaccine protects against infection, we expect cluster size will be reduced, as fewer students will be infected if they have been immunized. Observed cluster size may be reduced further even than this, if the vaccine allows harder-to-detect infections to occur. On the other hand, now more infectious variants of the coronavirus have emerged; the Alpha, Delta and Omicron variants have all had a higher estimated transmissibility than their predecessors (*8, 9*). Increased transmissibility would suggest larger cluster sizes, certainly among unvaccinated ages, but the relative impact of vaccination and the new variants together is difficult to gauge. Furthermore, changes in vaccination, transmission and immune evasion may all lead to a change in the variability in cluster sizes.

## Materials and Methods

Our data consist of reports of confirmed cases among students, teachers, and staff in schools in four Canadian provinces during the 2020–2021 school year. Data was collected by Dr Shraddha Pai with COVID Schools Canada (*6*), an initiative of the group Masks for Canada (*1*). We included the four provinces from this data set with the most schools reporting cases with date information. For each school, there is a list of confirmed cases among students, teachers and staff, along with the dates on which the cases were reported. We then assigned cases to clusters based on being at the same school and being reported within 7 days of each other; if the difference in date between two cases was less than or equal to 7 days, or they could be linked by a sequence of such cases, they were put in the same cluster. We chose 7 days on the basis of estimates of the serial interval for COVID-19 of approximately 5 days (*33*). Information was not available about whether the cases at the same school were in the same classroom. Accordingly, we interpret clusters as capturing all linked cases at a given school, and not just one classroom.

There is substantial uncertainty in whether each of our determined clusters of cases accurately represents a set of cases linked by transmission. For any cluster of two or more cases, it may be that two independent sets of cases are incorrectly included in the same cluster. This may lead us to overestimate the size of clusters. Likewise, any two of our clusters in the the same school that occur further apart than seven days apart may in fact be linked by a chain of undetected transmission, leading to an underestimate of cluster size. Both these factors may occur in our data, but we neglect both of them, taking the observed cluster size as given by our method. We are also unable to distinguish between transmission occurring in a school and in social activities with classmates outside of school.

In a given jurisdiction, we assume exposure events occur according to a Poisson process with variable rate. Independently of this process, once an exposure event occurs at a school, we say *Z* additional people are infected by the index case, for a total of *Z* + 1 individuals in the cluster. The variable *Z* includes individuals directly infected by the index case, as well as any subsequent infected individuals that are included in the same cluster. Following (*27*), we model *Z* as a Poisson random variable with parameter *ν*, where *ν* itself is a Gamma-distributed random variable. As described by (*27*), *Z* is then a negative binomial random variable. Rather than the usual parametrization of a negative binomial distribution we use parameters *R*_*c*_ and *k*. The parameter *R*_*c*_ is the expected number of additional infections in a cluster, and *k* is the dispersion: a measure of how far the distribution of *Z* is from being Poisson. As *k → ∞*, the distribution of *Z* approaches that of a Poisson distribution with mean *R*_*c*_. The variance of *Z* is *R*_*c*_(1 + *R*_*c*_*/k*) and so for smaller values of *k* we expect more of the secondary cases to occur in rare large clusters rather than in frequent small clusters (*27*).

There are then a total of *Z* + 1 infected individuals in the school. To give an idea of how the distribution of true cluster size depends on the parameters when they are in this range, in Figure 1 we show the theoretical distributions for varying parameters. On the left, we fix *R*_*c*_ + 1 = 2 and vary *k*. Decreasing *k* causes there to be more clusters of size 1 (i.e. no transmission) and more large clusters, but reduces the number of intermediate-sized clusters. On the right, we fix *k* = 0.3 and show the effect of varying mean cluster size *R*_*c*_ +1. As *R*_*c*_ increases, the frequency of clusters with no or little transmission decreases and the frequency of larger cluster sizes increases.

**Figure 1:**
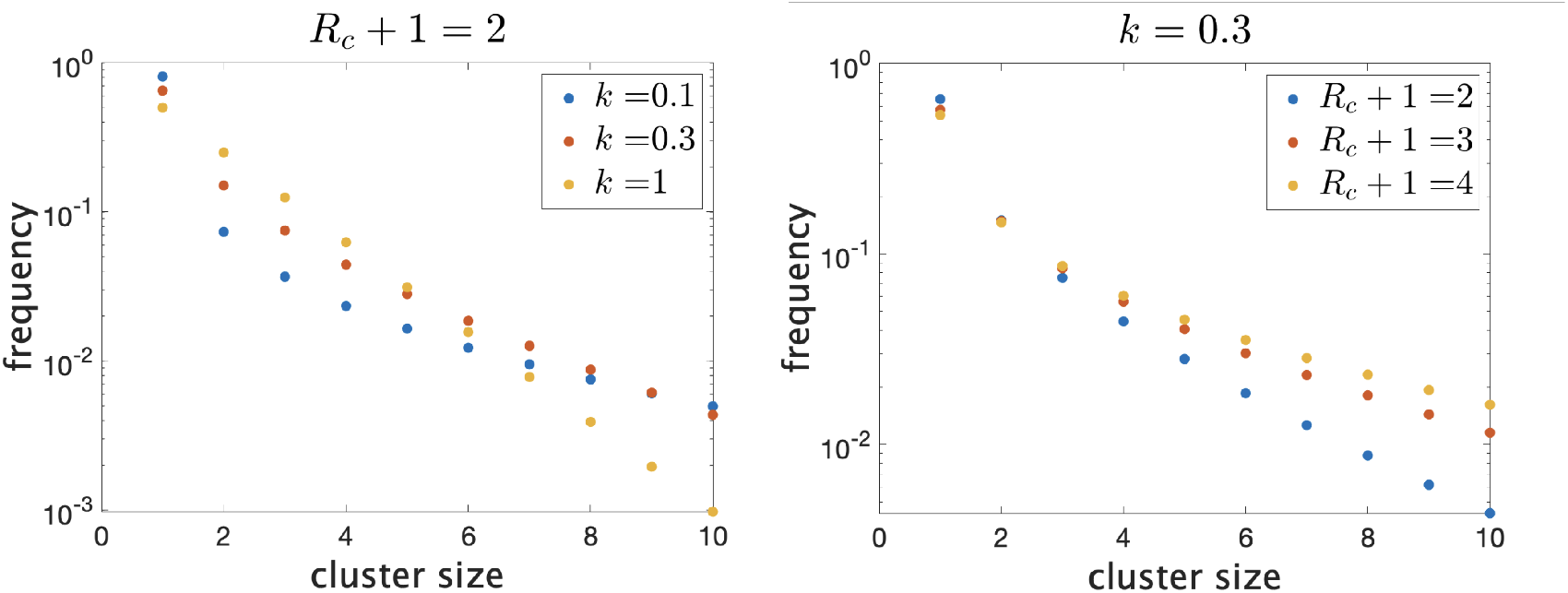
Frequency of clusters of different sizes on a log scale. Trends continue as shown for larger clusters. (left) Fixing mean cluster size *R*_*c*_ + 1 and varying dispersion *k*. (right) Fixing *k* and varying *R*_*c*_ + 1.

The number of the total *Z* + 1 cases that are actually observed, *X*, depends on the ascertainment model. We consider a model where each case is observed and contributes to the reported cluster size with probability *q*, so that the observed cluster size *X* (conditioned on *Z*) is binomial with parameters *n* = *Z* + 1 and probability *q*. The index case is treated the same as the infectees, so *X* may or may not include the index case. If none of the cases in a cluster are observed, we assume the cluster is not reported, so our model factors in the effect that smaller clusters are more likely to be missed. See the Supplementary Information for an explicit statement of the likelihood function.

For each collection of cluster sizes in our datasets we estimate the mean *R*_*c*_ and dispersion *k* using the ascertainment model with *q* = 0.75. We base this value on the meta-analysis (*5*) which reports ascertainment fractions for high-income regions in the Americas between 66% (in the last quarter of 2020) to 85% (in the second quarter of 2021). We use maximum likelihood estimation to obtain estimates of *R*_*c*_ and *k*, and we use the Hessian of the log-likelihood to obtain 95% confidence ellipses for the parameters (*45, Sec. 9.10*).

## Results

Figure 2 shows histograms of cluster size according to our definition in the four provinces. In Table 1 we show some statistics associated with the data for each province. In the top we show the number of clusters, the number of schools appearing, the number of schools with more than one reported cluster, and the fraction of schools with multiple clusters. In the bottom we show the fraction of clusters that have only one observed case, and the average number of observed cases in the clusters, the maximum observed cluster size, the index of dispersion (variance divided by mean) of cluster size, and index of dispersion of the number of cases in a cluster subtracting one for the presumed index case.

**Table 1:** (Top) For each of the four Canadian provinces: number of clusters in the data, number of schools reported, number of schools with multiple clusters, fraction of schools with multiple clusters. (Bottom) Fraction of clusters with one case, mean observed cluster size, maximum observed cluster size, and index of dispersion (variance of number of cases divided by mean number of cases) with and without subtracting one for the index case.

| Province | Number of Clusters | Number of Schools | Schools with Multiple Clusters | Fraction of Schools Multiple Clusters |
| --- | --- | --- | --- | --- |
| Manitoba | 1754 | 542 | 396 | 0.73 |
| Saskatchewan | 1211 | 466 | 295 | 0.63 |
| Ontario | 8482 | 3337 | 2147 | 0.64 |
| Alberta | 5032 | 1537 | 1158 | 0.75 |

Table 1: (Top) For each of the four Canadian provinces: number of clusters in the data, number of schools reported, number of schools with multiple clusters, fraction of schools with multiple clusters.
| Province | Fraction with One Case | Mean Observed Cluster Size | Max Observed Cluster Size | Index of Dispersion (IoD) | IoD without Index Case |
| --- | --- | --- | --- | --- | --- |
| Manitoba | 0.58 | 2.16 | 44 | 3.44 | 6.43 |
| Saskatchewan | 0.66 | 1.70 | 16 | 1.23 | 2.98 |
| Ontario | 0.63 | 1.83 | 50 | 1.87 | 4.13 |
| Alberta | 0.47 | 2.45 | 108 | 4.94 | 8.35 |

**Table 2:** Properties of the estimated distribution for the Poisson parameter *ν*, the index case-specific expected number of further cases in a cluster. The expected value of *ν* is *R*_*c*_ and its distribution gives important information about overdispersion of clusters. In units of hours^*-*1^.

| Province | Mean | Standard Deviation | Median | 90th percentile | 99th percentile |
| --- | --- | --- | --- | --- | --- |
| Manitoba | 1.43 | 2.45 | 4.3e-01 | 4.1 | 11.7 |
| Alberta | 1.86 | 2.55 | 8.9e-01 | 5.0 | 11.9 |
| Ontario | 1.04 | 1.70 | 3.5e-01 | 3.0 | 8.1 |
| Saskatchewan | 0.88 | 1.46 | 2.9e-01 | 2.5 | 7.0 |

**Figure 2:**
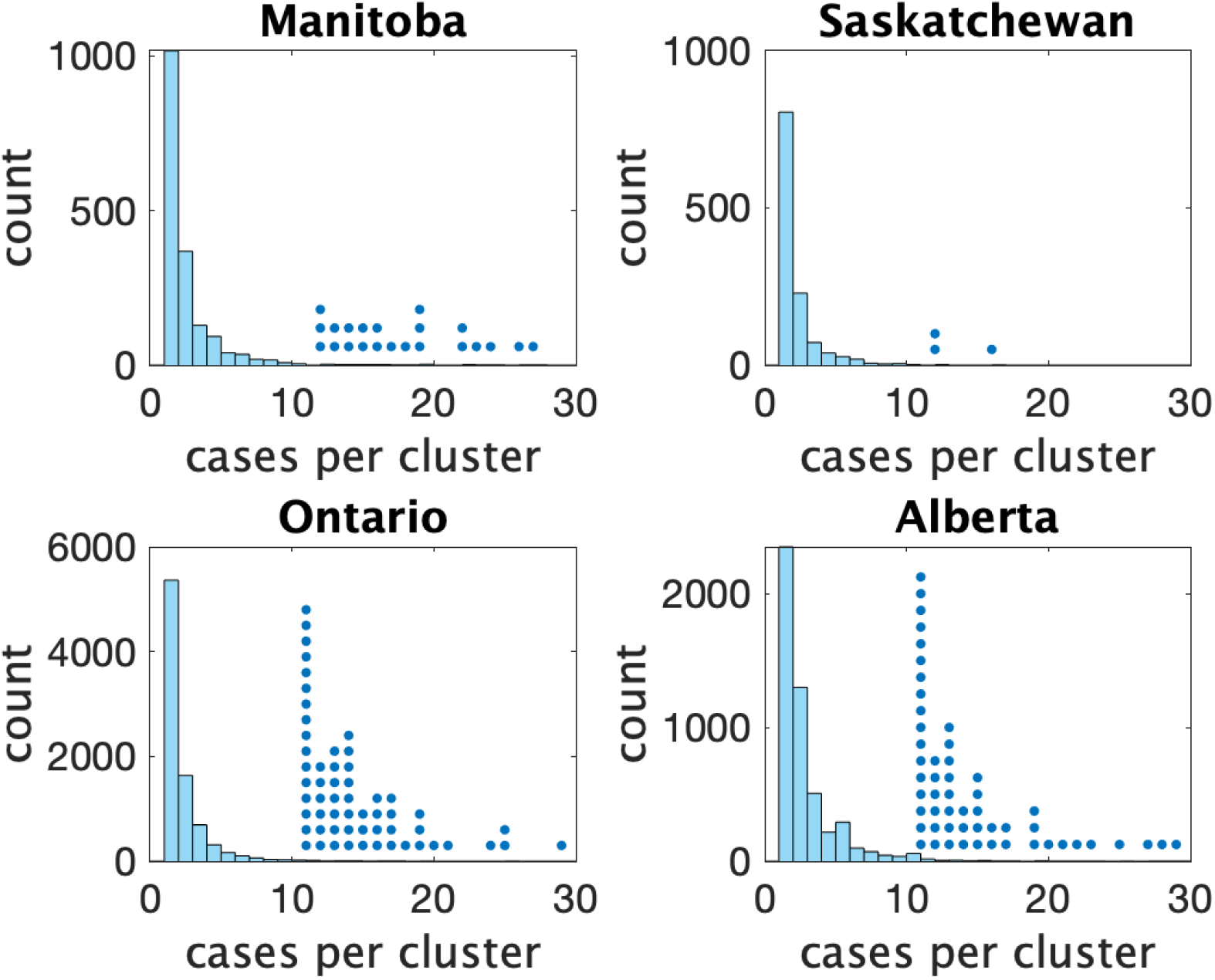
Histograms of observed cluster sizes in four Canadian provinces Each dot represents a single cluster of size 11 or larger, and indicates the presence of (more rare) larger clusters.

In Figure 3 (left) we show the rate (in clusters per day per 100,000 population) that cases appear in the dataset over time. In Figure 3 (right) we show the rate of COVID incidence per 100,000 population in the province over the same period of time. There is an apparent correspondence between the two time series, with peaks in rate of clusters per day occuring near peaks in incidence.

**Figure 3:**
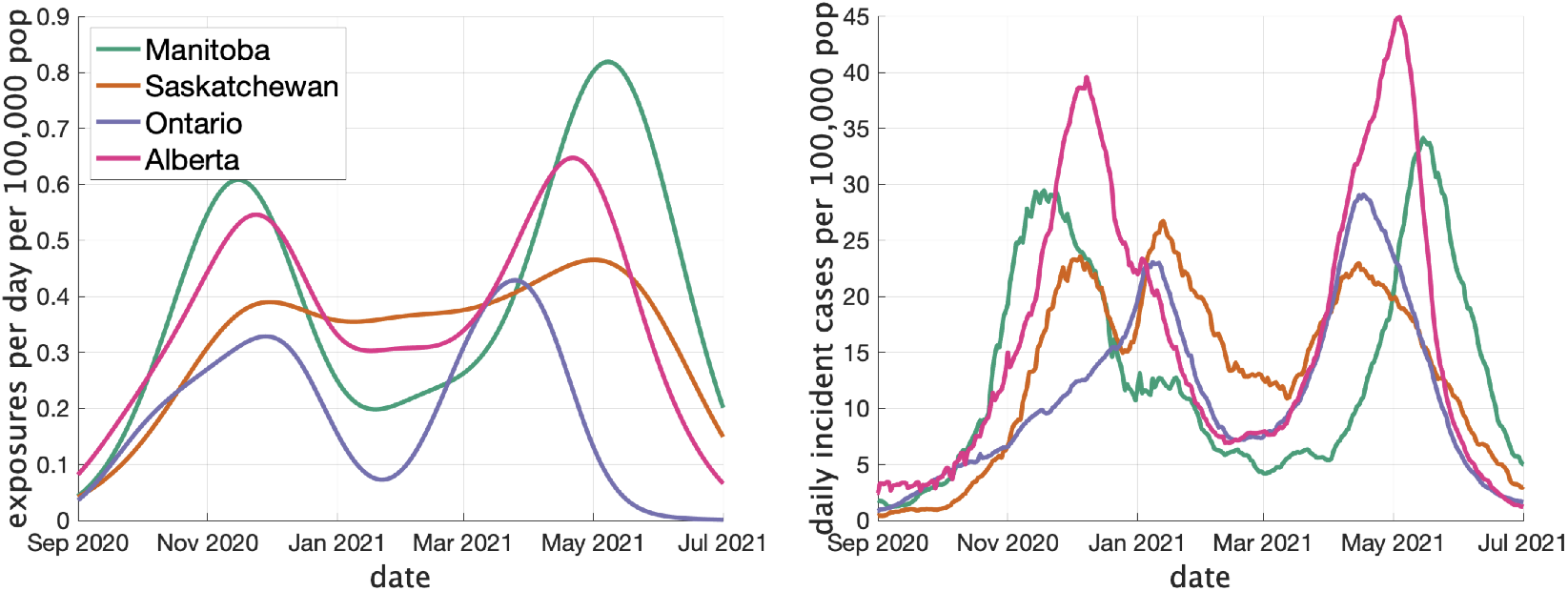
(Left) Estimates of the rate of new clusters (per 100,000 population) as a function of time in each province. (Right) Incident cases per day (per 100,000 population) in the same province over the corresponding time interval. Case counts are averaged over a two week window.

Figure 4 (right) shows the estimated mean cluster size (= *R*_*c*_ + 1) and dispersion *k* for the four Canadian provinces. Mean cluster sizes ranged from 1.9 to 2.9 cases, and dispersion ranged from 0.34 to 0.53 (recalling that no overdispersion corresponds to *k → ∞*.) Recall that we determined clusters by including cases in the same cluster if the were reported within 7 days of each other. If we extend this threshold to 10 days, mean cluster sizes increase modestly, ranging from 2.0 to 3.2.

**Figure 4:**
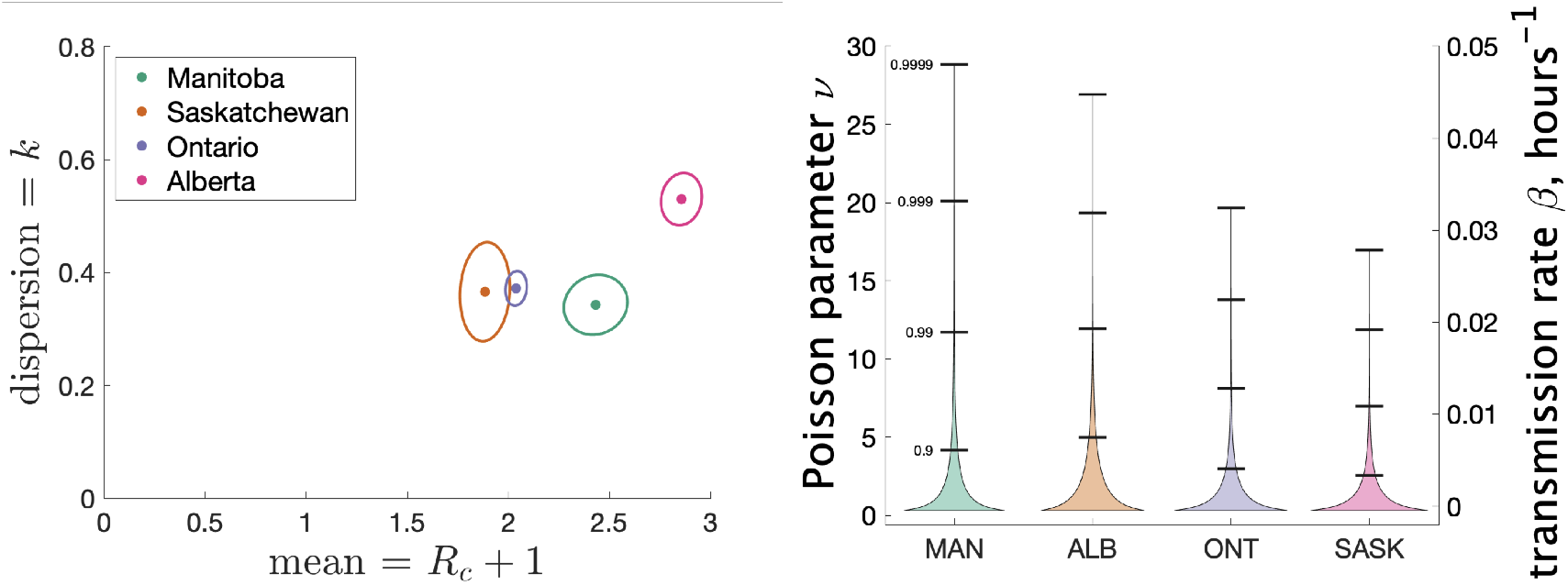
(left) Estimates of mean and dispersion of cluster size for four Canadian provinces using the individual ascertainment model with ascertainment rate 0.75. Estimate of mean includes index case. (right)Estimated distribution of *ν* (left axis) and instantaneous transmission rate *β* (right axis) for different provinces.

In the Supplementary Information we explore varying the ascertainment fraction between 0.2 and 1. Though lower ascertainment fractions yield bigger values of *R*_*c*_ and smaller values of *k*, we see that the parameter estimates are relatively insensitive to values of *q* between 0.5 and 1. For example, when *q*_1_ is reduced from 0.75 to 0.5, the range of *R*_*c*_ + 1 shifts from 1.9 – 2.9 to 3.2 – 6.4, and the range of *k* shifts from 0.34 – 0.53 to 0.22 – 0.39. The reason for this is that though a given cluster with multiple cases will look smaller with fewer cases detected, and lower detection will thereby bias observed size downwards, many single-case clusters will not be detected at all, biasing the observed cluster size upwards again. We also consider an alternate model of ascertainment, where the chance of a cluster being reported at all depends on the size of the cluster, and vary the rate of ascertainment in that alternate model; see the Supplementary Information.

As a way of interpreting dispersion values and what they mean for cluster size, we consider the fraction of all cases that occur in the largest 20% of all clusters. (If the distribution of cases follows the Pareto principle (*46*) then 80% of the cases will be in the top 20% largest clusters.) If we consider only secondary cases (not including the index case) we see from Figure 5 (right) the fraction that are due to the 20% largest clusters for various values of mean cluster size and *k*. For example, for Alberta with a mean cluster size of 2.9 and a dispersion *k* of 0.53, 69% of the secondary cases are in the top 20% of the clusters by size. For Saskatchewan, with a mean cluster size of 1.9 and *k* = 0.37, 82% of secondary cases are in the top 20% of clusters by size. When we include index cases, the fractions are correspondingly lower, as we see in Figure 5 (right).

**Figure 5:**
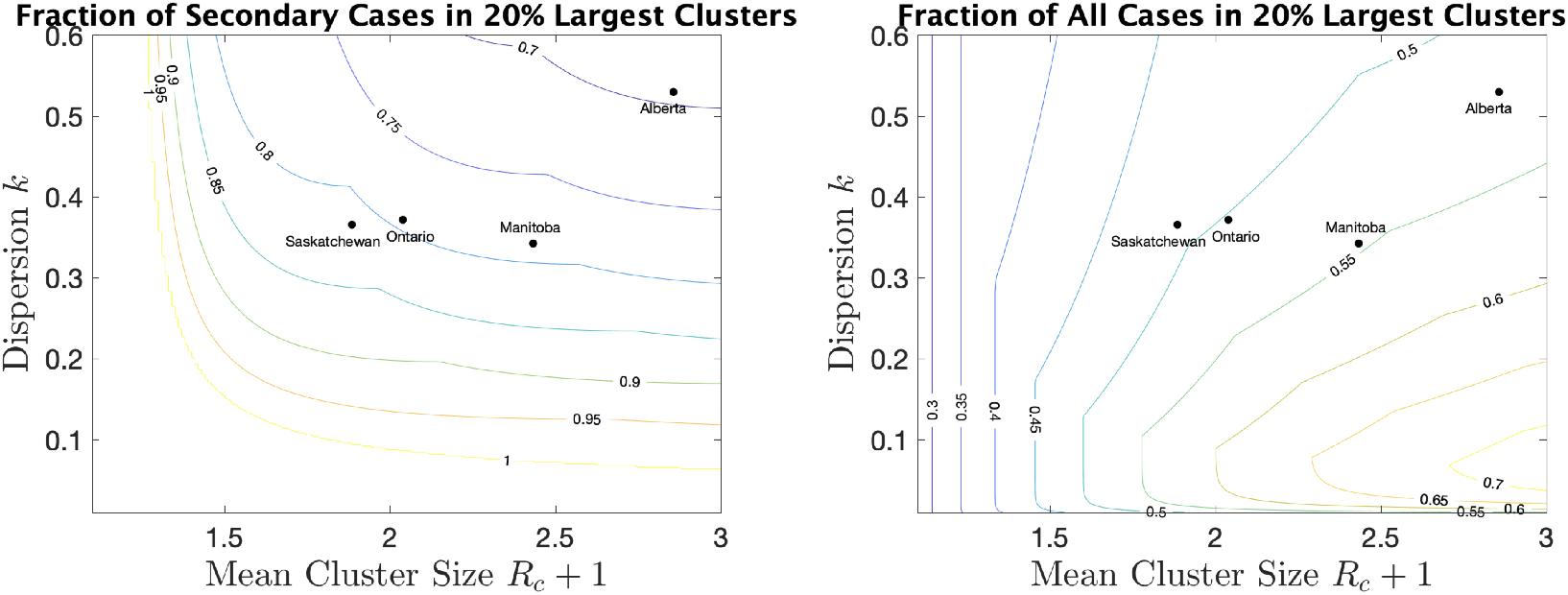
For a range of mean cluster size and dispersion *k*, the fraction of cases in the 20% largest clusters, counting only secondary cases (left), or all cases, index and secondary (right). Dots indicate the location of the four provinces in the plots.

Another way to visualize the variability of transmission we have inferred from the data is to show the distribution of the Poisson parameter *ν*, of which *R*_*c*_ is just the mean. In our model *ν* is the index case-specific expected number of further cases in a cluster, and is a gamma distributed random variable. Figure 4(right) shows the estimated distribution of *ν* for each jurisdiction, and Table 3 shows some key properties of the distribution for each of the provinces.

**Table 3:**
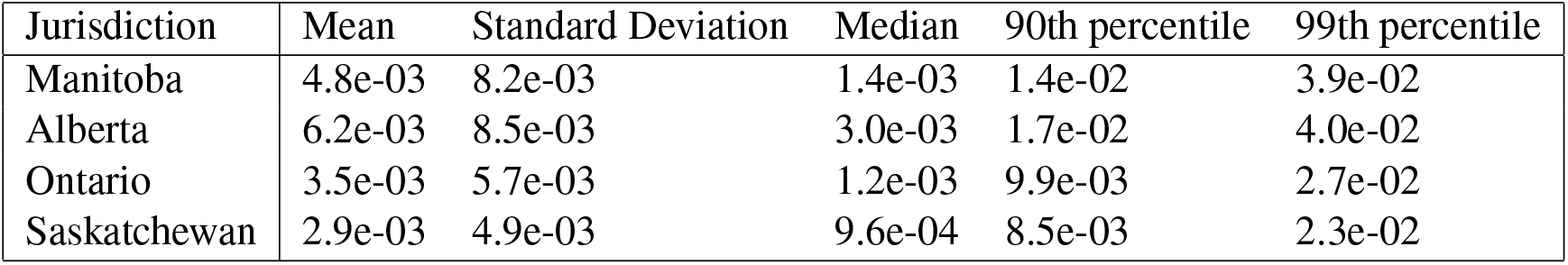
Properties of the estimated distribution for the instantaneous transmission rate *β*. In units of hours^*-*1^, with 95% credible intervals for the mean and standard deviation.

| Jurisdiction | Mean | Standard Deviation | Median | 90th percentile | 99th percentile |
| --- | --- | --- | --- | --- | --- |
| Manitoba | 4.8e-03 | 8.2e-03 | 1.4e-03 | 1.4e-02 | 3.9e-02 |
| Alberta | 6.2e-03 | 8.5e-03 | 3.0e-03 | 1.7e-02 | 4.0e-02 |
| Ontario | 3.5e-03 | 5.7e-03 | 1.2e-03 | 9.9e-03 | 2.7e-02 |
| Saskatchewan | 2.9e-03 | 4.9e-03 | 9.6e-04 | 8.5e-03 | 2.3e-02 |

Our model does not consider the details of transmission at the individual level, and so does not make use of an instantaneous transmission rate per contact pair. However, by making some simple assumptions about SARS-CoV-2 transmission, we can infer a distribution of transmission rate *β* from our estimate of the distribution of the parameter *ν*. Recall that *ν* is a Gamma-distributed random variable that gives mean number of secondary cases. Another way to estimate mean cluster size is to use an individual contact model where when an infectious person is in contact with a susceptible person, the susceptible person is infected with rate *β*. In such a model we assume that infected individuals are in a classroom for 2 days before isolating (when they develop symptoms), and that the total contact time with their classmates is *T* = 12 hours. Assuming that all individuals are in the same class, the infected individual is in contact with *n* = 25 other susceptible students for that time period. Then the infected individual will on average infect *βnT* other students. So we estimate *β* = *ν/*(*nT*). Since *ν* is Gamma-distributed, our estimate of *β* is too. Figure 4 (right) shows with the right *y*-axis the distribution of *β* for the different Canadian provinces. Table 3 shows some of the features of the estimated distribution for *β*.

One application of these estimates of the distribution of *β* is that we can explore the consequences of different types of interventions in the classroom setting. In (*40*) the authors consider a simple model of SARS-CoV-2 transmission among a group of contacts and investigate the quantity *R*_event_, the average number of secondary infections due to the presence of a single infectious individual. *R*_event_ is determined by *T*, the total length of time the infectious individual is with others; *n*_contact_, the number of contacts at any point in time, *τ* the length of time the individual is with a fixed set of contacts; and *β*, the instantaneous transmission rate. The parameter *τ* can vary between some fraction of *T* (for example *T/*3, if the index case divides their time equally between three sets of *n*_contact_ contacts) or *T* if the set of contacts is fixed. Interventions can be classified according to which of these parameters they modify: reducing transmission reduces *β*, social distancing reduces *n*_contact_, and “bubbling” (staying with the same small group rather than mingling) increases *τ* to *T*. If we use our distributions for *β* with the model of (*40*) we can estimate how the distribution of cluster sizes is changed with different interventions under different values of the parameters *R*_*c*_ and *k*.

In Figure 6 we show estimated size distributions of clusters under different interventions. Our baseline simulation settings intend to capture a pre-COVID high school classroom: *T* = 12 hours (two days of exposure before the index case isolates), *τ* = 3 hours (each student has four different classes that they attend for equal periods of time), *n*_class_ = 25, and *β* is sampled from our estimated distribution for a given choice of *R*_*c*_ and *k*. We consider three interventions: **transmission reduction** (for example, by introducing masks) reduces *β* by a factor of 2; **social distancing** cuts the size of a class in half; **strict bubbling** increases *τ* to *T*. For all values of *R*_*c*_ and *k* we consider, we simulate 10^7^ clusters to obtain a histogram of the number of secondary cases as well a mean and standard deviations, for the baseline conditions and for each of the three interventions, as shown in Figure 6. Means and standard deviations are accurate to the number of digits reported, and are shown with the corresponding histogram in the figure.

**Figure 6:**
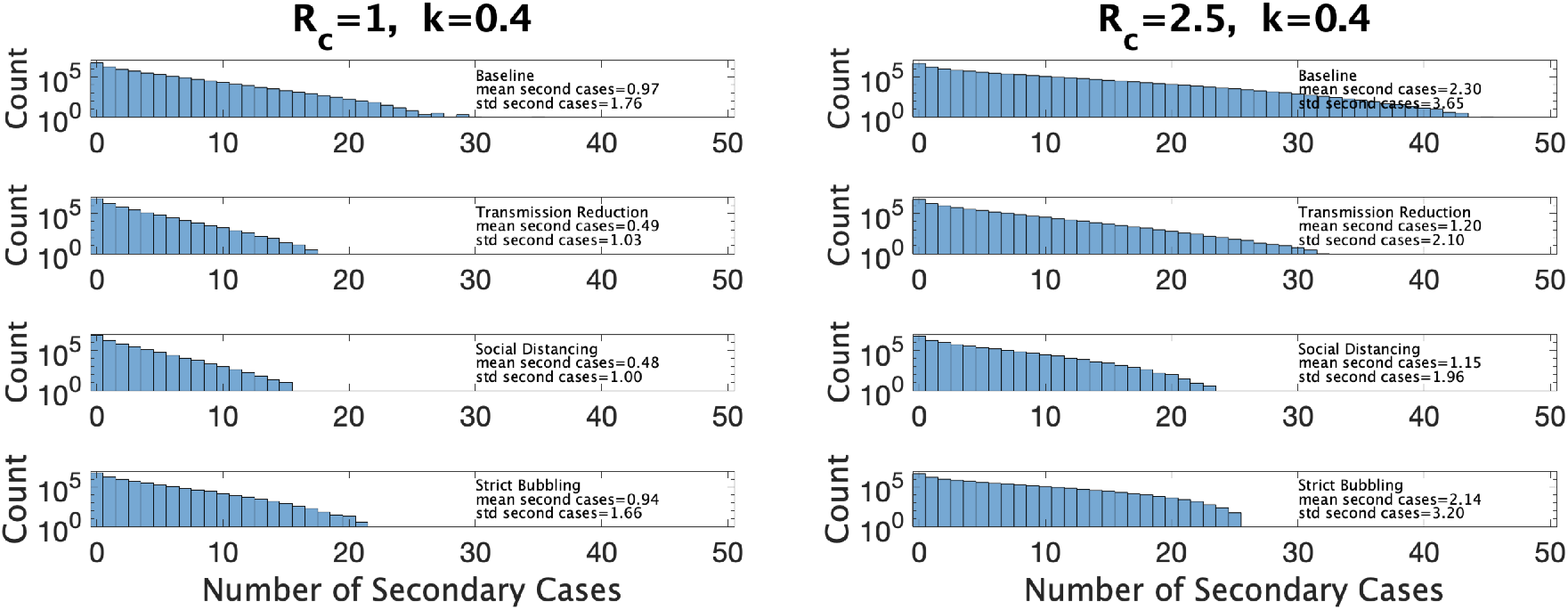
Distribution of the number of secondary infections under baseline conditions and under three interventions. Left: under parameter choice *R*_*c*_ = 1 and *k* = 0.4. Right: with *R*_*c*_ = 2.5.

Figure 6(left) shows results for *R*_*c*_ and *k* close to that of Ontario (*R*_*c*_ = 1.0, *k* = 0.4). We see that both reducing transmission and social distancing are effective in reducing the total number of cases, whereas bubbling does not contribute much to reduce cluster sizes. This is characteristic of what (*40*) call the linear regime: the number of secondary infections depends linearly on the time the infectious person is present with others. Figure 6 (right) shows the results in a hypothetical setting where *R*_*c*_ is much larger (*R*_*c*_ = 2.5, *k* = 0.4), perhaps due to the existence of a more transmissible variant such as Omicron. Here transmission reduction is less effective than in the linear regime, and strict bubbling more so; increasing *β* has moved us closer to the so-called saturating regime, where transmission reduction is relatively less effective than bubbling.

## Discussion

We have used cluster size data to estimate the mean and dispersion in cluster sizes, accounting for imperfect case detection. We have found that in each of the provinces we consider, the majority of school transmission occurs in a small number of classrooms, with the top 20% of clusters containing between 70% and 80% of the secondary cases in school settings. We developed a method to estimate the transmission rate per contact per unit time, with reference to a simple model of classroom transmission. Having a direct estimate of the transmission rate allows us to compare the benefits of different control measures. We find that with parameters estimated from Canadian jurisdictions during the 2020–2021 school year, interventions that reduce transmission rates (such as masking) and reduce number of contacts at any one time (class size reduction), are more effective than strategies aimed at keeping sets of contacts consistent (such as bubbling).

Overdispersion in transmission of SARS-CoV-2 and other infectious diseases is well documented (eg (*48*)) and is often described with reference to the 20/80 rule: that 20% of the infected individuals account for 80% of the transmission. Naturally, if the more infectious 20% can be identified, interventions targeting that portion of the population are likely to have a high impact. For SARS in 2003, Lloyd-Smith et al (*27*) estimated that 20% of the cases were responsible for almost 90% of the transmission. Estimates for SARS-CoV-2 also find considerable overdispersion, with the parameter *k* between 0.1 (*17*) and 0.5 (*24*) (with *R*_0_ = 2.5 this gives the top 20% of cases causing 69% - 96% of the transmissions; see for a survey). These estimates focus on the distribution of the number of people an infectious person infects directly during the whole course of infection (with mean *R*_0_), which is of obvious epidemiological importance, but for which it is difficult to obtain high-quality data. When a case is identified, we are not always able to determine who they infected, and indirect methods must be used. We may miss cases, and others may be wrongly attributed to a given index case.

In our present study, we examined a different random quantity, the number of additional cases *Z* infected, either directly or through intermediaries, by a given index case in a given setting. We denoted the mean of *Z* by *R*_*c*_. Including the index case means that the cluster size is *Z* + 1, with mean *R*_*c*_ + 1. Compared to estimates of *R*_0_, *R*_*c*_ does not count people infected at other sites, but it does include additional cases, because it includes both direct and indirect transmission. *Z* and its mean *R*_*c*_ are therefore more focused on the particular setting (in this case a school) than *R*_0_ is. In general it will depend on the infectiousness of the index case, as well as how conducive the environment is to transmission, and what activities are undertaken there. Determining the distribution of *Z*, as we have done here, provides an alternative means of investigating transmission.

However, these two measures of transmissibility (*R*_0_ and *R*_*c*_, the mean of *Z*) may be close enough that it is instructive to compare our estimates for *Z* with the traditional *R*_0_, and our dispersion estimates with dispersion estimates for the number of secondary infections. Our *R*_*c*_ ranges from 0.9 in Saskatchewan to 1.9 in Alberta. These low values of *R*_*c*_ are inconsistent with *R*_0_ estimates (which range from 2 to 6 (*3*)), and indicate that in these jurisdictions schools are unlikely to be a major contributor to SARS-CoV-2 spread. Our estimates for *k* range from 0.34 (in Manitoba) to 0.53 (in Alberta), corresponding closely to earlier estimates of dispersion.

Overdispersion has consequences for controlling transmission and for estimation. Estimating the average transmission rate from a small number of clusters will be difficult, and will result in a high variability. Most likely what will be observed in a small number of sampled clusters will be little to no onward transmission, which would lead to underestimates of the transmission rate. But if one or more larger clusters are included in a sample by chance, then this could lead to an overestimate of the transmission rate.

If we could identify the conditions under which the rare larger clusters occur (high-risk individuals, activities and settings) we could achieve a disproportionately large effect on reducing transmission by applying new measures in these settings. There are myriad possible reasons for overdispersion of transmission for SARS-CoV-2, including variation in viral load (*12*), behaviour, and number of contacts. But a key factor in higher dispersion with SARS-CoV-2 in comparison to other pathogens such as influenza is aerosolization (*19*), which allows the index case to infect others in the room even if they are not a close contact. Properties of the setting may be very important, with some settings (cramped, poor ventilation) being especially conducive to transmission. It would be good to identify classrooms or schools where there is a high risk of larger clusters. For example, if data were available on the occupancy, ventilation standards, mask use, classroom size, distancing behaviour and other features of classrooms, we could investigate how this related to the cluster size. Rapid tests may be especially good for identifying the most infectious individuals, given that they are sensitive to viral loads (*29*), but additional data collection is likely needed to quantify setting-level risks.

Two important changes have happened since the majority of the data here was collected. Firstly, in the jurisdictions studied, effective vaccines have been developed and deployed for those aged 512 and up (*38*). There are several ways in which this may effect cluster sizes in the school setting. To the extent that the general population (including adults) being vaccinated reduces incidence of COVID (*25,28,47*), there will be fewer introductions of SARS-CoV-2 into the classroom, and so fewer exposures will occur leading to fewer clusters. This effect may be dampened by relaxation of distancing and other measures that were keeping COVID-19 at bay and are no longer necessary in the context of vaccination. The distribution of cluster sizes when clusters do occur will also change: many students who might otherwise be infected will be protected by the vaccine, others who are vaccinated but infected (breakthrough infection) may have reduced symptoms and therefore may not be detected. We therefore expect the mean cluster size to be reduced by vaccination, in age ranges where vaccination has been deployed. It is unclear what the consequences will be for the dispersion.

Secondly, new, more transmissible variants of SARS-CoV-2 have emerged (*7*), most notably the Alpha variant, the Delta variant, the Omicron variant, and most recently the BA.2 strain of the Omicron variant, each with a substantially higher transmissibility than its predecessors. A natural way to implement this change in our model is to multiply *R*_*c*_ by an appropriate factor, boosting the size of clusters, without changing the dispersion parameter *k*. Data from the period in which Delta was the prevalent strain is not available, but schools in the Canada and the US saw resurgences in clusters in schools around school openings (*2, 15, 36*).

Our data and model have some limitations. The data rely on crowdsourcing, and there is reason to believe that reporting is incomplete. Inequity may effect data collection, as wealthier districts are more likely to have the resources to identify and track transmission. In general, larger clusters may be more likely to be reported. In the modelling, we assumed a Poisson random variable for the cluster size, with an underlying gamma-distributed rate variable. This is a flexible model allowing for considerable overdispersion, but it is simple and does not explicitly handle complexities such as the differences between elementary and high schools. Our estimates of the transmission rate were derived (where feasible) from a model with a fixed number of hours that the index case would be infectious in the classroom, and fixed class sizes. Accounting for variation in these would result in more variability in the estimates.

A major limitation of our analysis is how we assigned cases to clusters. Since the only data available was the number of cases reported on a given day at a school, we put cases in the same cluster if they occurred within 7 days of each other. The choice of 7 days was informed by the serial interval of COVID-19, but unavoidably, some cases will have been put in clusters that were not linked by transmission, whereas other that were linked were not put in the same cluster. Furthermore, we assumed that all clusters consisted of an index case and a number of other cases directly infected by the index case. In reality, there may be longer chains of transmission. Any of these assumptions may bias our estimates of the distribution of *ν* and *β*. Finally, our illustrative modelling of the impact of interventions was simple, and used simple assumptions for the impacts of masking, distancing and cohorts (bubbling). Our estimates of the per-contact transmission rate per unit time could, however, be used in more sophisticated simulation modelling to compare interventions.

Despite these limitations, our approach has distinct advantages. We have developed estimates of the person-to-person transmission rate derived directly from data. The data we use (cluster sizes) are relatively easy to access. This approach does not require individual-level data or contact tracing information, which are often not available; individuals may be identifiable and data are held within public health institutions. However, we note that if it were available, contact tracing data would be an excellent gold standard against which to check our assumptions about cluster identification. Our estimation approach, together with cluster size data, offers a high-resolution view of transmission: we can estimate the distribution of cluster sizes in specific settings, accounting for reporting and overdispersion, and in some contexts we can estimate the transmission rate, all without requiring either individual-level data or assumptions on transmission parameters such as the serial interval (see, in contrast, (*14, 43*) which require serial interval estimates). The results offer context-specific tools to simulate interventions in particular settings (here, schools). The methods are readily generalizable to other structured settings, such as workplace outbreaks where workplaces are similar in size and structure. Our results also suggest the need for data collection activities that can relate cluster sizes to setting variables such as occupancy, density, ventilation, activity and distancing behaviour. Ultimately this would provide the data needed to design interventions that best reduce school and/or workplace transmission.

## Data Availability

Data is all available at https://github.com/PaulFredTupper/covid-19-clusters-in-schools

## Acknowledgments

We thank Alisha Morris and the other volunteers at the National Education Association for providing the US data that was used in this study. PT and CC were supported by Natural Science and Engineering Research Council (Canada) Discovery Grants (RGPIN-2019-06911 and RGPIN-2019-06624). CC receives funding from the Federal Government of Canada’s Canada 150 Research Chair program.

### Appendix 1

#### Model for Cluster Size

We consider two models for ascertainment (whether a case is actually detected), though we only consider the first in the main text.

In the first ascertainment model (**Individual Ascertainment**) each of the infected individuals is detected with a probability *q*_1_. So *X*, the total number of infected individuals is binomial (*n, p*) with parameters *n* = *Z* + 1 and *p* = *q*_1_. If by chance none of the individuals are observed, we do not observe the cluster. This is meant to model a situation where cases are detected independently of each other, and one detected case does not lead to further tests or screening.

In the second ascertainment model (**Cluster Ascertainment**), at first each case is identified with probability *q*_2_, but then if any of the students are identified they are all subsequently identified. This is intended to capture a situation where a single detected case triggers testing for the whole class. Again, if no cases are detected we do not observe the cluster. This is equivalent to saying that clusters of size *m* are detected in their entirety with probability 1 *-* (1 *- q*_2_)^*m*^.

The number *Z* of new cases given the presence of one infectious case is a Poisson distributed random variable with a rate *ν* that is itself a Gamma-distributed random variable with a shape *k* and scale *θ*. This means *Z* has a negative binomial distribution NB(*r, p*) where *r* = *k* and *p* = *θ/*(1 + *θ*). Letting Θ = (*k, θ*), the pmf of *Z* is

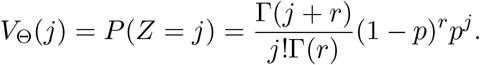

Under the individual ascertainment model with ascertainment probability *q*_1_, *X*, the number of observed cases, is binomial (*n, p*) with *n* = *Z* + 1 and *p* = *q*_1_. So, the probability that *i* individuals are observed is

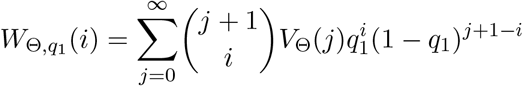

for *i* = 0, 1, Since we do not observe clusters with no observed cases the probability of observing a cluster of size *i* is actually 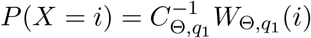 for *i* = 1, 2, …, where 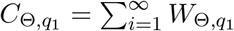.

If the observed cluster sizes are *X*_*i*_, *i* = 1, …, *n*, the log-likelihood function for Θ = (*k, θ*) under the individual ascertainment model is then

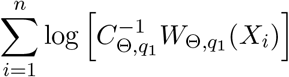

Under the cluster ascertainment model, the cluster is observed or not with probability 1*-*(1*-q*_2_)^*Y* +1^. So the probability of observing *X* = *j* in a cluster is

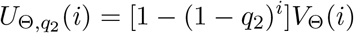

for *i* = 0, 1, … but then since we cannot observe clusters of size 0, an observed cluster has size *i* with probability

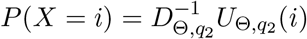

for *i* = 1, 2, …, where 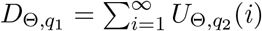.

If the observed cluster sizes are *X*_*i*_, *i* = 1, …, *n*, the log-likelihood function for Θ = (*k, θ*) under the cluster ascertainment model is then

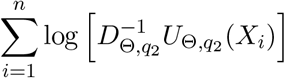

Under both ascertainment models, we then go from our estimates of *k* and *θ* to estimates of *R*_*c*_ via the formula

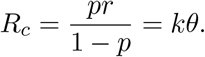

We use the Delta method to obtain confidence intervals for *R*_*c*_ from confidence intervals on *k* and *θ*.

#### Analysis of US Data

The US data was gathered from the National Educational Association website (*4*) (originally started by Alisha Morris, an educator at a Kansas high school) which collected data from news media and from reports submitted by volunteers (*42*). We selected the 8 states with the most data available, and covering the period between August and November 2020. For the US data we used confirmed student cases listed on a particular date for the cluster size, excluding teachers and staff. We did not collect cases reported on different days at the same school in the same cluster as we did with the Canadian data.

In Table 1 we show some statistics associated with the data for each state. In the top we show the number of clusters, the number of schools appearing, the number of schools with more than one reported cluster, and the fraction of schools with multiple clusters. In the bottom we show the fraction of clusters that have only one observed case, and the average number of observed cases in the clusters, the maximum observed cluster size, the index of dispersion (variance divided by mean) of cluster size, and index of dispersion of the number of cases in a cluster subtracting one for the presumed index case. Comparing with Table 1, we can see several striking differences between the US and Canadian data. There are substantially more clusters reported in Canada than in the US, despite the US states having greater population on average. This may partly be explained by the Canadian data being collected over a longer period than the US data, but this is likely not the full explanation: in Appendix 1 figure 2 we show the rate (in clusters per day) that cases appear in the dataset over time. We can compare with Figure 3 (left) that shows the same thing for the Canadian data. Even at times when both US and Canadian datasets record clusters, Canadian rates are higher than US rates by an order of magnitude, despite incidence rates being similar in US states versus Canadian provinces (Appendix 1 figure 2 (right) versus Figure 3 (right)). This suggests that the method used for gathering cluster reports in different jurisdictions varied substantially between the two datasets, especially when we look at daily incident cases in each. Furthermore, the majority of schools in the US datasets only report one cluster.

There are also substantial differences in statistics of cluster sizes. Mean observed cluster sizes were without exception larger in the US states than Canadian provinces, and Canadian provinces tended to have a higher fraction of clusters with only one case. Given the incomplete nature of the US data, we cannot determine whether these differences are due to real differences in transmission in the jurisdictions, or because smaller clusters were less likely to be reported in the US states.

**Table 1:** (Top) For each of the eight US states: number of clusters in the data, number of schools reported, number of schools with multiple clusters, fraction of schools with multiple clusters. (Bottom) Fraction of clusters with one case, mean observed cluster size, maximum observed cluster size, and index of dispersion (variance of number of cases divided by mean number of cases) with and without subtracting one for the index case.

| State | Number of Clusters | Number of Schools | Schools with Multiple Clusters | Fraction of Schools Multiple Clusters |
| --- | --- | --- | --- | --- |
| Texas | 369 | 326 | 30 | 0.09 |
| Florida | 147 | 134 | 10 | 0.07 |
| Ohio | 122 | 95 | 12 | 0.13 |
| Pennsylvania | 322 | 247 | 46 | 0.19 |
| Wisconsin | 130 | 118 | 11 | 0.09 |
| Georgia | 68 | 53 | 10 | 0.19 |
| Indiana | 105 | 84 | 13 | 0.15 |
| Illinois | 81 | 76 | 5 | 0.07 |

Table 1: (Top) For each of the eight US states: number of clusters in the data, number of schools reported, number of schools with multiple clusters, fraction of schools with multiple clusters.
| State | Fraction with One Case | Mean Observed Cluster Size | Max Observed Cluster Size | Index of Dispersion (IoD) | IoD without Index Case |
| --- | --- | --- | --- | --- | --- |
| Texas | 0.69 | 1.73 | 21 | 2.29 | 5.40 |
| Florida | 0.69 | 5.31 | 90 | 27.28 | 33.62 |
| Ohio | 0.69 | 2.11 | 21 | 4.30 | 8.18 |
| Pennsylvania | 0.73 | 1.75 | 28 | 3.84 | 8.95 |
| Wisconsin | 0.46 | 4.02 | 31 | 6.28 | 8.36 |
| Georgia | 0.46 | 4.76 | 38 | 10.09 | 12.77 |
| Indiana | 0.81 | 2.19 | 36 | 9.00 | 16.56 |
| Illinois | 0.52 | 3.47 | 36 | 11.07 | 15.55 |

Appendix 1 figure 3 shows the estimated mean cluster size (= *R*_*c*_ + 1) and dispersion *k* for the eight US states. In the US data, mean cluster size was estimated to range from about 2 in Texas to almost 8 in Florida. Dispersions ranged from 0.05 to 0.3, showing considerable overdispersion compared to the Poisson distribution. However, given that we are very probably substantially undersampling clusters in the US data, and the clusters that we are observing are likely larger ones, these estimates of mean cluster size are biased upwards in a way we are not able to control for.

**Appendix 1 - figure 1:**
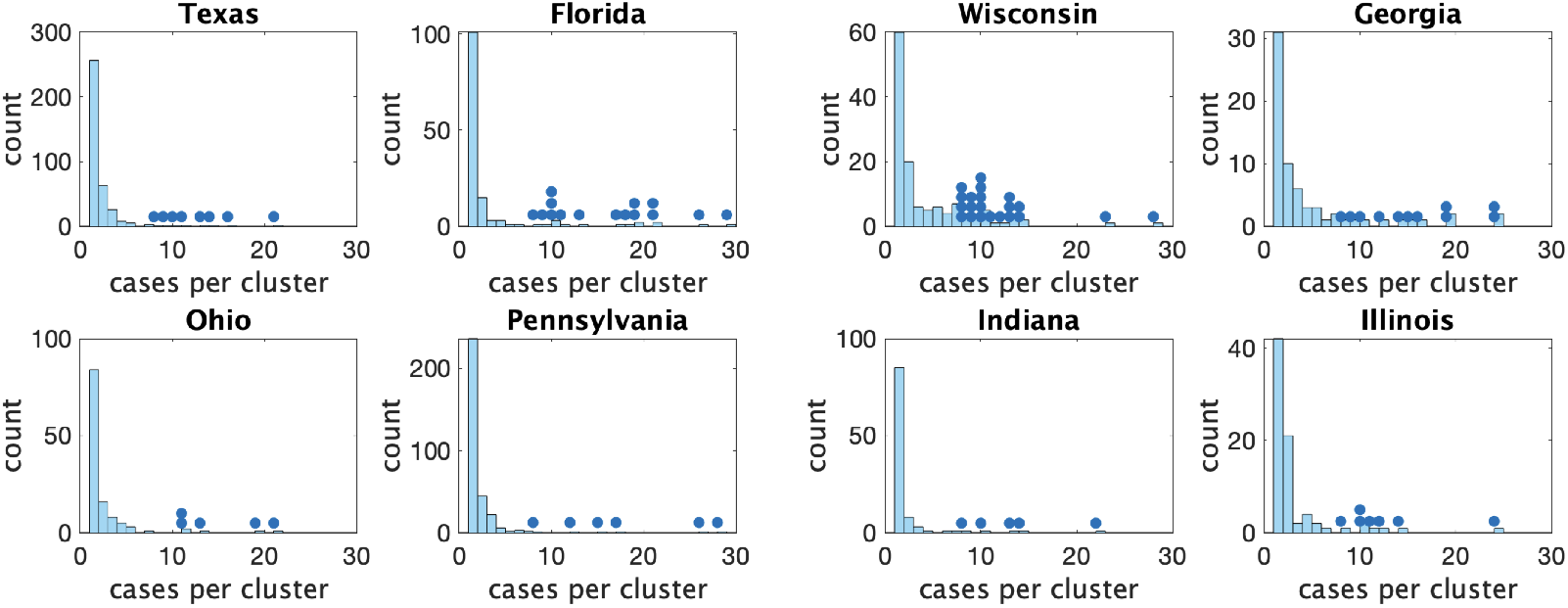
Histograms of observed cluster sizes in 8 US states We only show clusters of size 30 or fewer. Each dot represents a single cluster of size 8 or larger, and indicates the presence of (more rare) larger clusters.

**Appendix 1 - figure 2:**
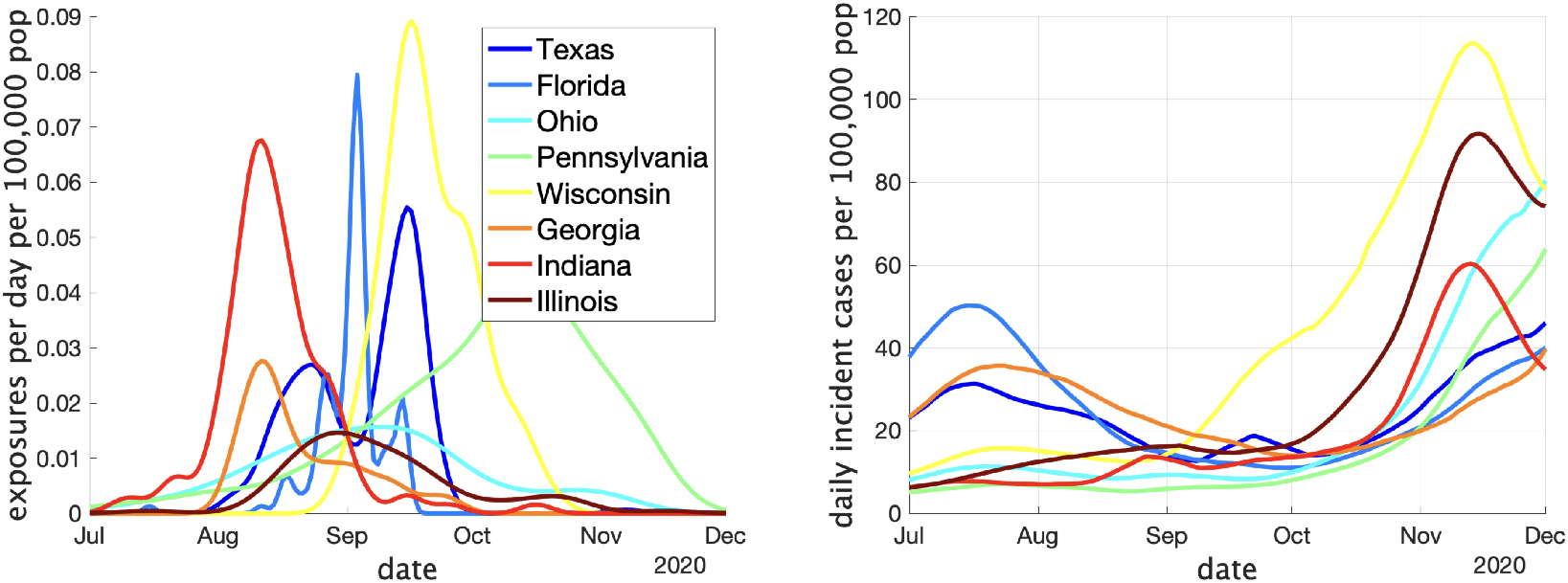
(Left) Estimates of the rate of new clusters being reported (per 100,000 population) as a function of time in each province. (Right) Incident cases per day (per 100,000 population) in the same province over the corresponding time interval. Case counts are averaged over a two week window.

**Appendix 1 - figure 3:**
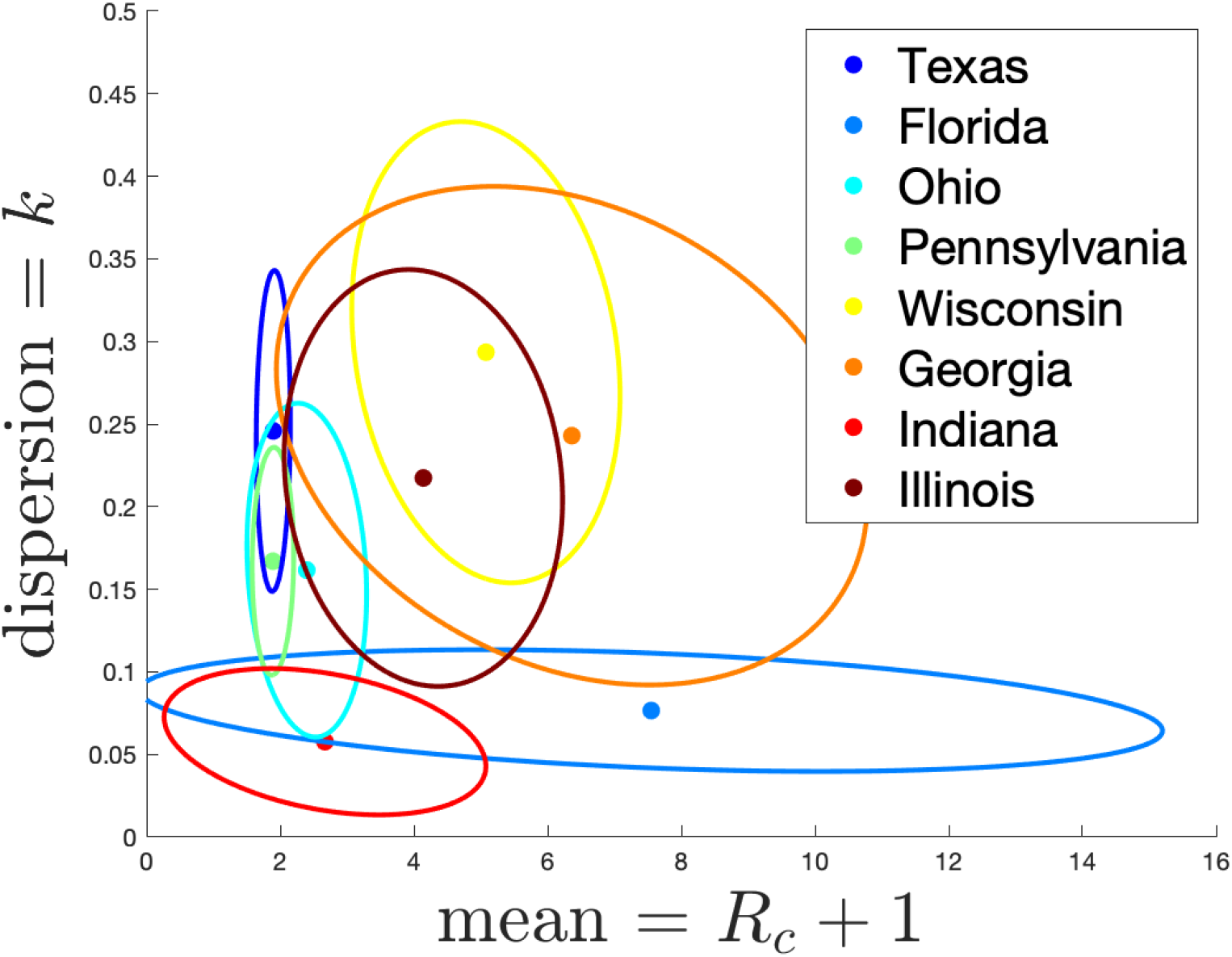
(left) Estimates of mean and dispersion of cluster size for eight American states using the individual ascertainment model with ascertainment rate 0.75. Estimate of mean includes index case.

#### Varying the rate and model of ascertainment

In the main text we estimated parameters with the assumption of the individual ascertainment model with an ascertainment probability of 0.75. Here we investigate how our main parameters *R*_*c*_ + 1 (expected cluster size) and *k* (dispersion) vary with this ascertainment probability. We also consider the alternate ascertainment model discussed in the previous section **Model for Cluster Size**.

Appendix 1 figure 4 shows the parameter estimates for the two models. The left plots show results for the individual ascertainment model where we set *q*_2_ = 1 and vary *q*_1_ from 0.2 to 1. The right plots show results for the group ascertainment model with *q*_1_ = 1 and *q*_2_ varying from 0.2 to 1. We see that the parameters do vary with the model and the ascertainment fraction, but relative magnitudes of the parameters in different jurisdictions do not change.

**Appendix 1 - figure 4:**
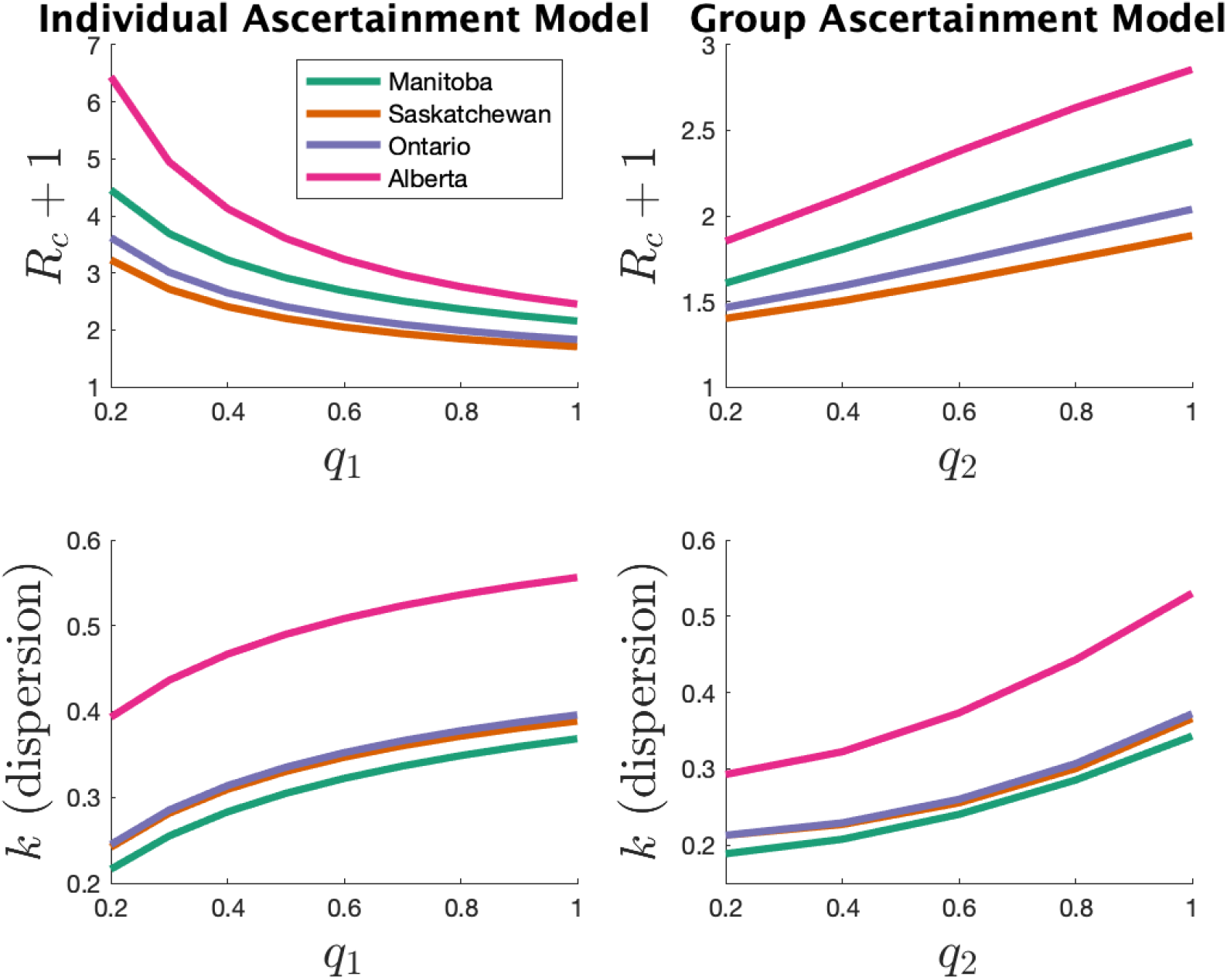
Estimates of mean and dispersion of cluster size for four Canadians provinces (right) using the individual ascertainment model (left) and the group ascertainment model (right) with varying ascertainment rate. Estimates of mean includes index case.

## References

1. Masks4Canada – canadian doctors, professionals, & citizens for masks. https://masks4canada.org/. Accessed: 2021-10-26.

2. Kids are facing covid-19 risks. here’s what parents can do - CNN video. CNN, August 2021.

3. Yousef Alimohamadi, Maryam Taghdir, and Mojtaba Sepandi. Estimate of the basic reproduction number for COVID-19: A systematic review and meta-analysis. J. Prev. Med. Public Health, 53(3):151–157, May 2020.

4. National Education Association. Nea school and campus covid-19 reporting site, https://app.smartsheet.com/b/publish?eqbct=00a2d3fbe4184e75b06f392fc66dca13, accessed: 12.04.2020.

5. Niklas Bobrovitz, Rahul Krishan Arora, Christian Cao, Emily Boucher, Michael Liu, Claire Donnici, Mercedes Yanes-Lane, Mairead Whelan, Sara Perlman-Arrow, Judy Chen, et al. Global seroprevalence of sars-cov-2 antibodies: A systematic review and meta-analysis. PloS one, 16(6):e0252617, 2021.

6. Covid Schools Canada. Canada covid-19 school tracker, https://covidschoolscanada.org/.

7. CDC. For parents: Multisystem inflammatory syndrome in children (MIS-C) associated with COVID-19. https://www.cdc.gov/mis/mis-c.html, September 2021. Accessed: 2021-10-26.

8. CDC. SARS-CoV-2 variant classifications and definitions. https://www.cdc.gov/coronavirus/2019-ncov/variants/variant-info.html, December 2021. Accessed: 2022-4-11.

9. CDC. Omicron variant: What you need to know. https://www.cdc.gov/coronavirus/2019-ncov/variants/omicron-variant.html, March 2022. Accessed: 2022-4-11.

10. Centers for disease control and prevention. Science brief: Transmission of sars-cov-2 in k-12 schools and early care and education programs – updated. https://www.cdc.gov/coronavirus/2019-ncov/science/science-briefs/transmission k 12 schools.html, September 2021. Accessed: 2021-10-26.

11. Sonia Chaabane, Sathyanarayanan Doraiswamy, Karima Chaabna, Ravinder Mamtani, and Sohaila Cheema. The impact of COVID-19 school closure on child and adolescent health: A rapid systematic review. Children, 8(5), May 2021.

12. Paul Z Chen, Niklas Bobrovitz, Zahra Premji, Marion Koopmans, David N Fisman, and Frank X Gu. Heterogeneity in transmissibility and shedding SARS-CoV-2 via droplets and aerosols. October 2020.

13. Victor Chernozhukov, Hiroyuki Kasahara, and Paul Schrimpf. The association of opening K-12 schools with the spread of COVID-19 in the united states: County-level panel data analysis. Proc. Natl. Acad. Sci. U. S. A., 118(42), October 2021.

14. Anne Cori, Neil M Ferguson, Christophe Fraser, and Simon Cauchemez. A new framework and software to estimate time-varying reproduction numbers during epidemics. Am. J. Epidemiol., 178(9):1505–1512, November 2013.

15. Beth Reese Cravey. Duval schools report almost 1,500 COVID-19 cases; 2 schools remain closed due to outbreaks. Florida Times-Union, August 2021.

16. Itai Dattner, Yair Goldberg, Guy Katriel, Rami Yaari, Nurit Gal, Yoav Miron, Arnona Ziv, Rivka Sheffer, Yoram Hamo, and Amit Huppert. The role of children in the spread of COVID-19: Using household data from bnei brak, israel, to estimate the relative susceptibility and infectivity of children. PLoS Comput. Biol., 17(2):e1008559, February 2021.

17. Akira Endo, Centre for the Mathematical Modelling of Infectious Diseases COVID-19 Working Group, Sam Abbott, Adam J Kucharski, and Sebastian Funk. Estimating the overdispersion in COVID-19 transmission using outbreak sizes outside China. Wellcome Open Res, 5:67, July 2020.

18. Ontario Government. Schools COVID-19 data - ontario data catalogue. https://data.ontario.ca/dataset/summary-of-cases-in-schools. Accessed: 2021-10-27.

19. Ashish Goyal, Daniel B Reeves, E Fabian Cardozo-Ojeda, Joshua T Schiffer, and Bryan T Mayer. Viral load and contact heterogeneity predict sars-cov-2 transmission and super-spreading events. Elife, 10:e63537, 2021.

20. Douglas N Harris. When should schools reopen fully in-person? https://www.brookings.edu/blog/brown-center-chalkboard/2020/09/29/when-should-schools-reopen-fully-in-person/, September 2020. Accessed: 2021-10-26.

21. Fraser Health. School exposures. https://www.fraserhealth.ca/schoolexposures. Accessed: 2021-10-26.

22. Vancouver Coastal Health. COVID-19 potential exposure events in VCH schools. http://www.vch.ca/covid-19/school-exposures. Accessed: 2021-10-27.

23. Rebecca L Laws, Rebecca J Chancey, Elizabeth M Rabold, Victoria T Chu, Nathaniel M Lewis, Mark Fajans, Hannah E Reses, Lindsey M Duca, Patrick Dawson, Erin E Conners, Radhika Gharpure, Sherry Yin, Sean Buono, Mary Pomeroy, Anna R Yousaf, Daniel Owusu, Ashutosh Wadhwa, Eric Pevzner, Katherine A Battey, Henry Njuguna, Victoria L Fields, Phillip Salvatore, Michelle O’Hegarty, Jeni Vuong, Christopher J Gregory, Michelle Banks, Jared Rispens, Elizabeth Dietrich, Perrine Marcenac, Almea Matanock, Ian Pray, Ryan Westergaard, Trivikram Dasu, Sanjib Bhattacharyya, Ann Christiansen, Lindsey Page, Angela Dunn, Robyn Atkinson-Dunn, Kim Christensen, Tair Kiphibane, Sarah Willardson, Garrett Fox, Dongni Ye, Scott A Nabity, Alison Binder, Brandi D Freeman, Sandra Lester, Lisa Mills, Natalie Thornburg, Aron J Hall, Alicia M Fry, Jacqueline E Tate, Cuc H Tran, and Hannah L Kirking. Symptoms and transmission of SARS-CoV-2 among children - utah and wisconsin, March-May 2020. Pediatrics, 147(1), January 2021.

24. Ramanan Laxminarayan, Brian Wahl, Shankar Reddy Dudala, K Gopal, Chandra Mohan, S Neelima, K S Jawahar Reddy, J Radhakrishnan, and Joseph A Lewnard. Epidemiology and transmission dynamics of COVID-19 in two Indian states. Science, September 2020.

25. Eyal Leshem and Annelies Wilder-Smith. COVID-19 vaccine impact in israel and a way out of the pandemic. Lancet, 397(10287):1783–1785, May 2021.

26. Andrew T Levin, William P Hanage, Nana Owusu-Boaitey, Kensington B Cochran, Seamus P Walsh, and Gideon Meyerowitz-Katz. Assessing the age specificity of infection fatality rates for COVID-19: systematic review, meta-analysis, and public policy implications. Eur. J. Epidemiol., 35(12):1123–1138, December 2020.

27. James O Lloyd-Smith, Sebastian J Schreiber, P Ekkehard Kopp, and Wayne M Getz. Superspreading and the effect of individual variation on disease emergence. Nature, 438(7066):355–359, 2005.

28. Smriti Mallapaty. China COVID vaccine reports mixed results - what does that mean for the pandemic? Nature, January 2021.

29. Michael J Mina and Steven Phillips. Opinion. The New York Times, October 2021.

30. British Columbia Ministry of Education. K-12 education restart plan. https://www2.gov.bc.ca/assets/gov/education/administration/kindergarten-to-grade-12/safe-caring-orderly/k-12-education-restart-plan.pdf, July 2020. Accessed: 2021-10-26.

31. State of Michigan. School-Related cluster and outbreak reporting. https://www.michigan.gov/coronavirus/0,9753,7-406-98163_98173_102480---,00.html. Accessed: 2021-10-27.

32. Bellwether Education Partners. Missing in the margins 2021: Revisiting the COVID-19 attendance crisis. https://bellwethereducation.org/publication/missing-margins-estimating-scale-covid-19-attendance-crisis, October 2021. Accessed: 2021-10-26.

33. Balram Rai, Anandi Shukla, and Laxmi Kant Dwivedi. Estimates of serial interval for covid-19: A systematic review and meta-analysis. Clinical epidemiology and global health, 9:157–161, 2021.

34. Mendel E. Singer, Ira B. Taub, and David C Kaelber. Risk of myocarditis from covid-19 infection in people under age 20: A population-based analysis. medRxiv, 2021.

35. Kim Sneppen, Bjarke Frost Nielsen, Robert J. Taylor, and Lone Simonsen. Overdispersion in covid-19 increases the effectiveness of limiting nonrepetitive contacts for transmission control. Proceedings of the National Academy of Sciences, 118(14), 2021.

36. Star staff and wire services. Today’s coronavirus news: Ontario reports 748 cases; province still in fourth wave as cases rise, says ontario’s top doctor. The Toronto Star, November 2021.

37. Carole H Sudre, Benjamin Murray, Thomas Varsavsky, Mark S Graham, Rose S Penfold, Ruth C Bowyer, Joan Capdevila Pujol, Kerstin Klaser, Michela Antonelli, Liane S Canas, Erika Molteni, Marc Modat, M Jorge Cardoso, Anna May, Sajaysurya Ganesh, Richard Davies, Long H Nguyen, David A Drew, Christina M Astley, Amit D Joshi, Jordi Merino, Neli Tsereteli, Tove Fall, Maria F Gomez, Emma L Duncan, Cristina Menni, Frances M K Williams, Paul W Franks, Andrew T Chan, Jonathan Wolf, Sebastien Ourselin, Tim Spector, and Claire J Steves. Attributes and predictors of long COVID. Nat. Med., 27(4):626–631, April 2021.

38. The New York Times. See how vaccinations are going in your county and state. The New York Times, December 2020.

39. Zeynep Tufekci. This overlooked variable is the key to the pandemic. Atl. Mon., September 2020.

40. Paul Tupper, Himani Boury, Madi Yerlanov, and Caroline Colijn. Event-specific interventions to minimize COVID-19 transmission. Proc. Natl. Acad. Sci. U. S. A., 117(50):32038–32045, December 2020.

41. Russell M Viner, Oliver T Mytton, Chris Bonell, G J Melendez-Torres, Joseph Ward, Lee Hudson, Claire Waddington, James Thomas, Simon Russell, Fiona van der Klis, Archana Koirala, Shamez Ladhani, Jasmina Panovska-Griffiths, Nicholas G Davies, Robert Booy, and Rosalind M Eggo. Susceptibility to SARS-CoV-2 infection among children and adolescents compared with adults: A systematic review and meta-analysis. JAMA Pediatr., 175(2):143–156, February 2021.

42. Tim Walker. Teacher creates national database tracking COVID-19 outbreaks in schools. https://www.nea.org/advocating-for-change/new-from-nea/teacher-creates-national-database-tracking-covid-19-outbreaks. Accessed: 2021-10-27.

43. Jacco Wallinga and Peter Teunis. Different epidemic curves for severe acute respiratory syndrome reveal similar impacts of control measures. Am. J. Epidemiol., 160(6):509–516, September 2004.

44. Sebastian Walsh, Avirup Chowdhury, Vickie Braithwaite, Simon Russell, Jack Michael Birch, Joseph L Ward, Claire Waddington, Carol Brayne, Chris Bonell, Russell M Viner, and Oliver T Mytton. Do school closures and school reopenings affect community transmission of covid-19? a systematic review of observational studies. BMJ Open, 11(8), 2021.

45. Larry Wasserman. All of statistics: a concise course in statistical inference. Springer Science & Business Media, 2013.

46. Wikipedia contributors. Pareto principle. https://en.wikipedia.org/w/index.php?title=Pareto-principle&oldid=1052101893, October 2021. Accessed: NA-NA-NA.

47. Annelies Wilder-Smith and Kim Mulholland. Effectiveness of an inactivated SARS-CoV-2 vaccine. N. Engl. J. Med., 385(10):946–948, September 2021.

48. M E Woolhouse, C Dye, J F Etard, T Smith, J D Charlwood, G P Garnett, P Hagan, J L Hii, P D Ndhlovu, R J Quinnell, C H Watts, S K Chandiwana, and R M Anderson. Heterogeneities in the transmission of infectious agents: implications for the design of control programs. Proc. Natl. Acad. Sci. U. S. A., 94(1):338–342, January 1997.

